# Perspectives on multimorbidity care provision among public hospital-based healthcare workers in Blantyre and Chiradzulu, Malawi: A qualitative study

**DOI:** 10.1101/2025.06.06.25329117

**Authors:** Gift Treighcy Banda-Mtaula, Ibrahim G. Simiyu, Sangwani Nkhana Salimu, Stephen A Spencer, Nateiya M. Yongolo, Marlen S. Chawani, Hendry Sawe, Jamie Rylance, Ben Morton, Adamson S. Muula, Eve Worrall, Felix Limbani, Miriam Taegtmeyer, Rhona Mijumbi, the Multilink consortium

**Author notes:** Corresponding author: Treighcy, (GTB). Joint Senior author (Felix Limbani, Miriam Taegtmeyer, Rhona Mijumbi). Membership of the Multilink Consortium is provided in the Acknowledgements.

## Abstract

Multimorbidity, the presence of multiple chronic health conditions, is a leading cause of death globally. In Malawi, chronic noncommunicable and communicable diseases such as HIV frequently co-exist, putting pressure on an under-resourced system. However, the health system is primarily structured around disease-specific (vertical) programs, which hinders person-centred care approaches to multimorbidity. Our study focuses on multimorbidity care and explores the perceptions of healthcare workers on the patient pathways and service organisation throughout the patient’s interaction with the health facilities.

This cross-sectional qualitative study took an interpretivist approach. We conducted 13 days of clinical observations at Queen Elizabeth Central Hospital and Chiradzulu District Hospital. We also conducted 13 days of clinical observations and semi-structured in-depth interviews with different cadres of purposively sampled healthcare workers (n=22) at Queen Elizabeth Central Hospital and Chiradzulu District Hospital. Through thematic analysis, we identified an understanding of the organisation of care and healthcare workers’ perspectives on the delivery of services.

Findings showed both hospitals provided services for inpatients and outpatients with multimorbidity, including screening, management, prevention of secondary conditions and rehabilitation. Patient diagnosis and management for multimorbidity were often delayed due to frequent stockouts of medication and consumables necessary for diagnostic testing for NCDs at the hospital level. Some healthcare workers were not equipped with the knowledge, skills, or guidelines to manage multimorbidity. As HIV care is currently better resourced than other chronic conditions, healthcare facilities may strengthen the supply chain, healthcare workers’ training sessions and monitoring and evaluation tools to ensure NCDs are well managed, learning from HIV programmes.

## Introduction

Multimorbidity, the presence of multiple chronic conditions, is a stressor to health systems leading to poor quality of life, disability, treatment complications, and mortality.(1–3) In sub-Saharan Africa (sSA), Roomaney *et al* and Kasambara *et al* report a rising double burden of chronic communicable such as the human immunodeficiency virus (HIV) and non-communicable diseases (NCDs; e.g. hypertension). In Malawi, the health system is heavily dependent on donor funding for vertical (single-disease focused) health programmes, which leads to fragmented care for people with multimorbidity, that is inefficient and costly to the health system.(4)

The World Health Organization (WHO) advocates for person-centred care focusing on individuals needs, values and preferences and integrated, holistic disease management for people living with multimorbidity.(5) The Malawi Ministry of Health have demonstrated a commitment to these aims through its policies, including the Health Sector Strategic Plan III 2023-2030, which calls for integration across all levels of health care.(6) However, implementation is uneven and often relies on external funders. Partners in Health (PIH), an international nonprofit global health organisation, supports the Malawi Ministry of Health in implementing comprehensive integrated human immunodeficiency virus (HIV) and NCDs care in integrated chronic care clinics (IC3 model) in some primary and secondary healthcare facilities. (7) Integrated service delivery has been reported to improve retention to care,(8) improve clinical outcomes, and is considered to be economically feasible for Malawi.(9) Yet, health systems and clinical guidelines that are primarily designed for individual diseases, hinder healthcare providers’ ability to provide integrated management of multimorbidity.(10, 11) This can also lead to a high prevalence of poorly controlled chronic diseases in hospital emergency departments as well as at community level. (12, 13)

Previous qualitative studies relating to multimorbidity have largely focused on patient and caregivers’ perspectives and experiences of living with multimorbidity(14–16) or perspectives of both healthcare workers and patients, with a focus on patient experience.(17–19) However, healthcare workers’ perspectives have been understudied. Yousif *et al.* argued that the perspectives of healthcare workers are important because they are the primary providers of care and are key in influencing the success of prescribed care delivery and pathways of care within facilities. (20)This study draws on direct clinical observations of patient management and healthcare worker perspectives of care to understand how health services are organised for adults living with multimorbidity in Malawi. We seek to answer the question, “What are the perspectives of healthcare workers towards how clinical services are organised for patients with multimorbidity who present to the hospitals?” We defined multimorbidity as the presence of two or more of HIV, type 2 diabetes, hypertension and chronic kidney disease.

-c

## Methods

This study is embedded within a multi-country prospective cohort study undertaken as part of the Multilink programme.(21) Multilink aims to provide a comprehensive description of multimorbidity within secondary care in Africa, and subsequently to design and evaluate a complex intervention which identifies patients with multimorbidity during emergency assessment in hospitals in Malawi and Tanzania, optimising immediate treatment and ensuring post-discharge linkage to appropriate care.(22)

### Study design

Based on our research question, we sought to understand how services are organised for adults living with multimorbidity, and healthcare workers perspectives and experiences on providing care. Therefore, this was a cross-sectional qualitative study that adopted an interpretivist approach.(23) We collected data through clinical observations and in-depth interviews with healthcare workers with the aim of integrating the two data sources to get an in-depth understanding.

### Study Setting

We conducted this study in Malawi, with a population of 22 million people. Healthcare, at point of service, is free at government-owned facilities. The study was conducted in two such public hospitals in the Southern part of Malawi: 1) Queen Elizabeth Central (QECH) hospital in Blantyre, which is the largest national referral hospital (1,350 bed capacity), 2) Chiradzulu District Hospital (Chiradzulu), a secondary facility (300 bed-capacity) located 22.5km from Blantyre which refers complex patients to QECH. Both sites are expected to provide multimorbidity screening, treatment and follow up.

### Study participants and recruitment for in-depth interviews

We purposively sampled different cadres of healthcare workers who manage patients’ medical conditions in the Accidents and Emergency Department (QECH), outpatient department (Chiradzulu) and medical wards (both sites). We recruited consultants (medical officers with specialised training), medical officers (medical degree), clinical officers (diploma in clinical medicine), nurses (degree or diploma in nursing) and pharmacists in order to ensure representation of seniority, gender balance and a balance of facility types. We only recruited licensed healthcare workers with at least 3 months post-license work experience. Participant recruitment was based on convenience and continued until no new information emerged (data saturation)

### Data collection and management

We collected data from 17 January to 23 August 2023. The lead author (GTB, a PhD candidate trained in qualitative research methods) conducted 13 days of semi-structured observations over 4 weeks, following an agreed structured observation guide (Sup 1). The purpose was to iterate topic guides, understand the patient’s pathway and gain familiarity with the hospital context and get to know hospital staff. We observed patient activities as they walked in and met a healthcare worker. We then conveniently selected patients in the medical waiting area to observe their consultation sessions with healthcare workers to understand the interaction between healthcare workers. Notes captured patient interactions, processes, how and when patients undergo registration, triage, and procedures for attending to patients that have been admitted; patient management, and availability of resources.(24) During clinical observations, we wrote down patient contact points. Clarifications were sought from the healthcare workers and patients being observed when required for further contextual understanding of the processes. GTB wrote observation summaries at the end of each observation session. GTB and FL identified and discussed the themes from the observation summaries.

We developed an initial topic guide for in-depth interviews based on the components of the WHO health systems building blocks, concentrating on the service delivery building block (Sup 1).(25) We then used the emerging themes from the observation summaries to expand the interview topic guide before piloting. GTB conducted semi-structured interviews face-to-face in a private room within the hospital facilities. Interviews lasted 30 minutes to an hour. The interviews by GTB were conducted in English, with participants mixing some Chichewa phrases. GTB is fluent in both languages. Interviews were audio-recorded, transcribed and translated into English where necessary. GTB and FL (also bilingual and fluent in English and Chichewa)listened independently to the audio recording and matched English transcripts to ensure quality.

### Data analysis

We conducted thematic analysis approach as described by Braun and Clarke.(26) Briefly, GTB, IS, and MT independently read transcripts to familiarise themselves with the data and, coded the first three transcripts in parallel before developing a coding framework. Once the codebook was finalised, GTB coded transcripts in NVIVO 12 software.(27) We constantly juxtaposed findings from observations and interviews to understand the organisation of care and contextual aspects to reach a more comprehensive understanding. The analysis employed both deductive and inductive driven by our interest in the healthcare workers’ perspectives. We then charted the data to see where patterns converged or diverged and grouped codes to come up with emerging themes and sub-themes. We also incorporated data from various cadres and validated the quotes with study participants to ensure that they were quoted in the right context.(28)

### Reflexivity

The study team comprised clinical and non-clinical members, which increased the chances of understanding the data in multiple ways. The lead author completed her internship as a physiotherapist at Queen Elizabeth Central Hospital, which made her familiar with some of the clinical processes, staff and facilitated data collection. GTB undertook an objective approach, following the semi-structured observation and topic guides. We also incorporated data from various cadres and validated the quotes with study participants to ensure that they were quoted in the right context.(28) We have reported this study in line with the standard for reporting qualitative research guidelines (Sup 2).(29)

### Ethics Statement

The study received ethical approval from the College Medicine Research & Ethics Committee (COMREC) (Malawi)Ref P.11/21/3462 and the Liverpool School of Tropical Medicine Ref. 21-086 (UK). All participants in our study provided written consent.

## Results

We followed eight individual patients’ journeys from entry until they were discharged or admitted and allocated a bed. We conducted 22 interviews with healthcare workers with varying work experience, including the eight who were part of the observations during structured observations (Table 1). One healthcare worker at Chiradzulu and three at QECH who were approached, declined to participate in our study. These were replaced with consenting healthcare workers.

**Table 1:**
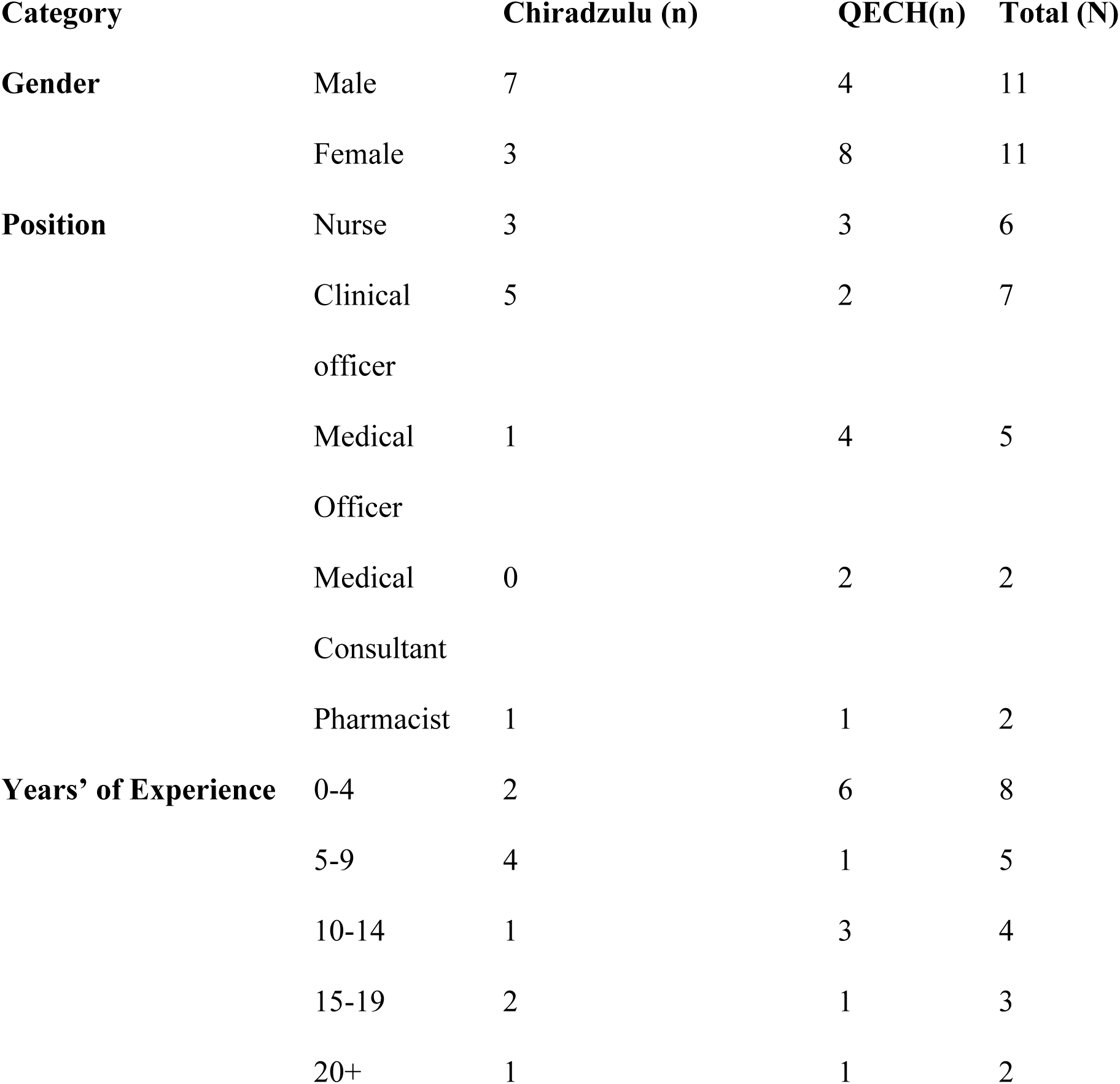
Characteristics of study participants (N=22)

Findings are grouped into five major themes: available services for adults living with multimorbidity, Complex care needs for patients, provider capacity to manage multimorbidity, institutional capacity to manage multimorbidity, and recommendations to improve care. Each theme is divided into subthemes that further explain the perspectives of healthcare workers.

### Theme 1:Available services for adults living with multimorbidity

Both sites provide multidisciplinary inpatient and outpatient care. Outpatient care includes scheduled outpatient clinics. Figure 1 illustrates patient interaction between services and the scheduling of outpatient clinics. We noted the patient pathway for multimorbidity was the same as for general medical patients within each facility until the time to allocate patients to outpatient clinics.

**Fig 1:**
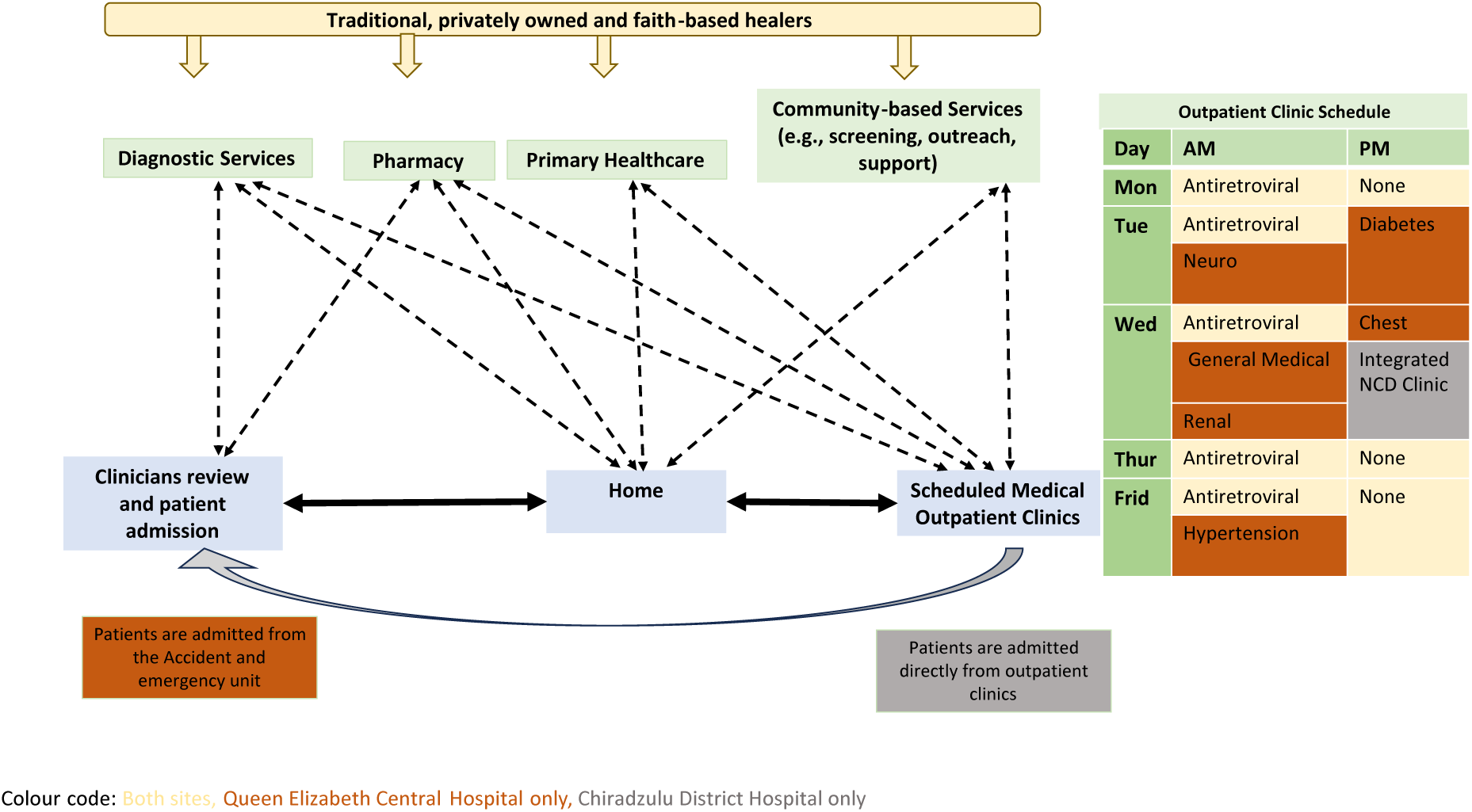
The organisation of care for patients with multimorbidity linking to long-term outpatient clinics at Chiradzulu and Queen Elizabeth Central Hospital.

### Routine outpatient clinics

Clinicians reported managing and monitoring stable patients through routine outpatient clinics. Chiradzulu had once weekly integrated NCD clinic and a daily Antiretroviral therapy (ART) clinic. However, the NCD clinic only catered to hypertension and diabetes. Other NCDs apart from cancer (seen in palliative care clinics) are seen as general outpatient cases. Clinicians reported patients were screened for NCDs at ART clinics but were referred to NCD clinics for management. Since the outpatient clinics started, clinicians at both sites acknowledged the increasing number of patients seen at these clinics.

*“Initially we were booking about 50 patients but our clinics are still being overwhelmed so, we are actually increased the numbers.”* (QECH_HCW05,female,10 years’ experience)

“…*currently, the numbers (of patients with multimorbidity) are going up and you can easily miss one or two things.*” (Chiradzulu,female,10 years’ experience)

At QECH, outpatient clinics for people not living with HIV were disease-specific, occurring at different days and times(Fig 1). Patients with multimorbidity either go to multiple clinics or one clinic, at the discretion of the clinician. QECH offered daily integrated HIV+NCD clinics for stable patients operated by the Lighthouse Trust (a centre of excellence in HIV management).

“*Lighthouse is a game changer for HIV, because they get everything done there. If they have other conditions, then they are managed there unless if the patient needs admission or if they need specialist care, then they’ll send them (to specialist clinics).” (QECH,HCW09,male, 4 years’ experience)*

Clinicians reported noting frequent occurrence of HIV with hypertension and diabetes, and diabetes and hypertension. They acknowledged patients living with multiple chronic conditions required services from multiple disciplines that were not always in sync. They reported booking times depended on the clinician’s perception of the acuity or severity of the condition.

*“So, on discharge, you‘re booked a clinic for review and referred to all the departments that they think you could benefit from, for example, Physiotherapy. But you are not guaranteed to get all services on the same day, sometimes you wait for the next available slot.” (QECH, HCW06, male, 6 years’ experience)*

At both sites, outpatient clinics outside of HIV (with special staff) were ran by the same clinicians who also attended to inpatients on clinic days. Because of their multiple duties, clinicians believed scheduling clinics made workload management easier. Some clinicians found weekly outpatient clinics inadequate to cater to the burden of multimorbidity and others found them inconvenient and not well coordinated between and among departments led to uncoordinated care and multiple hospital visits by the patient.

*“Patients come in the morning for fasting blood glucose check and wait for the clinic at 1:30pm. They spend the whole day here. The pharmacy closes at 4pm even on clinic days. You might be seen in the clinic, but find the pharmacy closed, meaning you collect your medication the next day.”* (QECH, HCW11, female,4years’experience)

During clinics, person-centered care was attempted in terms of identifying patient needs, but was reported as inadequate and sometimes not feasible due to increased workload

*“The (integrated NCD) clinic has more than 50 patients. They need to have vital signs and sugar levels checked, which take time. When they go to a clinician, a full assessment takes time, and other people wait for a long time to be seen. So we do not ask every patient their personal preferences.”* (Chiradzulu,HCW13,female,13 years’ experience)

Yes I can say we swap but its not like really swapping but when you have another issues on that day so you cannot attend the clinic so the staff can join you unto the clinic most of the time but there is other person for that but they are other providers which are assigned to do that but maybe due to other maybe attitude or what or what, so we can say like that.”

Clinicians reported patients that needed admission from the outpatient clinics were directly sent to the wards at Chiradzulu, while they were referred to the accident and emergency and trauma (AETC) at QECH. Clinicians at QECH reported that admitting outpatients from the clinics through the accident and emergency department means patients queue twice, which prolongs the admission process and inconveniences patients.

*“Patients from the diabetic clinic are told to go to AETC for admissions, yet they are the same clinicians who will be admitting them. So, we have these complications that are there that cause unnecessary delays” (QECH, HCW 01,female, 10 years’ experience)*

*“And there are instances where patients will end up going home because they are frustrated from being sent back and forth. They’ll be sitting (queueing) for an hour until somebody discovers they’re coming from the outpatient clinic. They already have a plan. What they need is admission”(QECH,HCW09,male,4 years’ experience)*

QECH has dedicated rooms for outpatient clinics located near the hospital entrance but away from the point of entry, which clinicians found conducive. At Chiradzulu, clinicians were concerned the infrastructure was not conducive for integrated clinics because the waiting area becomes crowded, which made patients uncomfortable.

“*It’s not very convenient because there is a combination of patients attending the clinic and the general OPD, so there is disorganisation and lack of privacy. People don’t feel like they are at their own clinic*” (Chiradzulu, HCW16, Male, 5years’experience)

### Value for integrated chronic disease management

There were conflicting views on the importance of integration for multimorbidity care. Most healthcare workers at Chiradzulu reported integrated out-patient clinics are better and improve the skills of the clinicians, whilst most clinicians at QECH found the concept of a specialised clinic to be better than one integrated clinic because clinicians get to focus on something they’re experienced in and, gain advanced specialist knowledge.

*“Combined clinics are much better than having patients seeing different specialists because the condition can be managed at once, so monitoring them is easy and you [the clinician] also get better.” (*Chiradzulu, HCW13,female, 10 years’ experience)

*“We have generalists looking at those conditions and they do their best according to the general medicine knowledge that they have, but there are just certain nuances or certain small details for a patient who has complicated things that a specialist better manages.”* (QECH, HCW01, female, 10years’experience)

### Continuation of care and patient follow-up

At both sites, clinicians reported lack of follow-up of patients who missed outpatient clinic appointments. The ART clinic follows up with patients in the community because they have sufficient funds for healthcare workers to phone patients, while that option does not exist for NCDs which leaves NCD patients’ attendance unmonitored.

*“I’ve never followed up (patients with NCDs), I don’t even know who hasn’t showed up. I can only check their file and be like oh you were supposed to come on this date written here, how come you didn’t?”* (QECH, HCW02,female, 3years’experience)

Clinicians reported counselling was an important and routine thing before each clinic started and involved patients and their caregivers. Counselling included health promotion, drug adherence, dieting, physical activity, and general mental health and well-being. They also reported encouraging patients to self-test to monitor their health at home. Self-testing was seen an unattainable for some patients.

*“We guide them on doing physical exercises and self-tests. We encourage them to have their own glucometers and BP machines. But not everyone can buy the equipment.”* (Chiradzulu, HCW15, male,4years’experience)

All clinicians sometimes perform occasional (unscheduled) community outreach to screen and monitor patients with chronic conditions. They also referred patients to primary facilities, either for routine tests in cases where they could not afford self-tests, or drug refills if the medication were available at the primary facilities. Some clinicians felt healthcare workers at the primary level did not have the capacity or resources to manage multimorbidity. This led to patients presenting to hospitals in an acute state because of delayed diagnosis or misdiagnoses at the primary care level.

“*Sometimes a patient may present late to a hospital, not because she was not serious about her illness, it could be because she has been to various facilities (primary healthcare) and was misdiagnosed.” (QECH, HCW11, male, 25 years’ experience)*

### Theme 2: Complex care needs for patients

Our clinicians acknowledged multimorbidity care was complicated due to disease-disease interaction, which increased the risk of more morbidities.

*“Diabetes, hypertension and HIV predispose you to renal impairment and put you at increased risk of other comorbid conditions, including heart disease. As a clinician, you have to be aware of and screen for.” (QECH,HCW08,female,4 years’ experience)*

Some felt the need for multiple medications was sometimes unavoidable in multimorbidity, which inconveniences patients.

*“Patients don’t like it, but if somebody has diabetes, hypertension, had a stroke and they also have arthritis, you can’t run away from giving them too many drugs. We try our best to limit the list to make the drugs twice a day. It’s not always possible to prescribe that way.”* ((QECH, HCW01, female, 10 years’ experience)

Clinicians felt the interaction of multiple diseases required more skill and knowledge, especially when it came to prescribing medication to avoid potential drug-drug interactions which some reported aren’t covered in guidelines.

*“Drug to drug interaction is challenging because you really need to think and the guidelines can’t cover everything. So with the background of the medications, you know which drugs interact though it’s not in the guideline.”*(Chiradzulu,HCW18,female,13 years’ experience)

Clinicians felt that the chronicity and life-long drug use was difficult for patients to comprehend and accept. In such cases, they felt patients either stopped taking their medication or resulted to herbal medicine in search for a cure.

*“People don’t understand that they will be taking the medication all their life, so they prefer to get it treated and done with. That’s what the herbal medications offer, it says that you will get treated once you take it. So, they would prefer that.”* (QECH, HCW02,female, 3years’experience)

Chronic diseases often require lifestyle changes, especially diet changes, which seen as difficult to maintain for some patients due to broader socioeconomic factors.

*“We advise them on what kind of food to eat and avoid; But not everyone can afford to change their meals.”* (Chiradzulu, HCW21, male,4years’experience)

### Theme 3: Provider capacity and support to manage multimorbidity

#### Lack of senior support and limited communication

Some clinicians felt unprepared to manage multimorbidity and reported lacking support in making clinical decision. Clinicians reported clinicians at QECH and Chiradzulu have a shared WhatsApp group where clinicians from Chiradzulu can consult remotely. Considering this is done on mobile devices, the clinicians felt feedback was not always timely as the QECH clinicians are busy during work hours. Even some clinicians at QECH felt that seniors or subspecialists *“are not always available” t*o respond to queries and can be hard to reach. The unavailability of seniors was attributed to a shortage of staff.

*“If a patient has hypertension and chronic kidney disease and we need the renal team to consult. It’s not easy for them to come maybe because there’s only one (specialist). I think that’s a challenge.”* (QECH, HCW05,female, 10years’experience)

Besides improving supervision and increasing the number of clinicians, some healthcare workers felt communication between the different multidisciplinary team could be improved, as in-patient departments still operated in siloes.

“*I think we need to improve the mode of communication. We don’t have the doctors’ rota so sometimes you come for night duty, you don’t know who to call to come and see the patient.”* (QECH, HCW07, female, 2years’experience)

### The need for on-the-job-training and refresher courses

There was a clear imbalance between knowledge of the management of HIV and NCDs. Most of the lower cadres of clinical workers attributed this to a lack of on-the-job training for NCDs whilst receiving several updates for HIV care. Some reported the occasional continuous professional development (CPD) sessions are normally disease-specific, which makes the management of multimorbidity a challenge. Some clinicians felt ill-equipped to manage the complexity (managing regimens, disease-disease interactions) of multimorbidity without further training. Some clinicians felt training needs to go beyond first-line care providers and extend to other non-clinical staff such as data clerks because they also play a role in patient management.

*“ I think we need training in managing patients with multimorbidity. Just like they train people in managing HIV patients, if there is a new guideline they train so we need the same in NCDs especially because we are seeing a lot of NCDS these days. So, all the nurses, all the clinicians need to be trained, as well as the data clerks when collecting the data need to be trained.” (Chiradzulu, HCW16, male, 5 years’ experience)*

### Theme 4: Institutional capacity to manage multimorbidity

#### Availability of staff

At both sites, we observed most service providers were nursing students. Our clinicians reported staff shortages (availability on site) which affected service delivery and contributed to long waiting times.

*“For example at the district hospital we have got few medical officers and they are busy people at the DHO. At the same time they are the ones with advanced knowledge (than clinical officers) so it becomes a challenge with us when maybe you are seeking for second opinion on how best you can manage this patient”*(Chiradzulu, HCW22,male,25 years’ experience)

Some nurses and medical officers reported increased workload and experiencing burnout, which led to limited consultation time with patients and affected quality of care.

#### Availability of diagnostic equipment and medication

Healthcare workers expressed challenges in diagnosing and managing conditions outside of HIV, due to limited availability of diagnostic equipment. Both facilities reported frequent breakdowns of diagnostic machines, missing reagents and frequent stock out of test kits, such as glucose sticks and batteries for blood pressure cuffs. During observations, we also noted a patient whose blood glucose couldn’t be measured because the glucometer was missing.

Clinicians at both facilities appreciated the supply of antiretroviral therapy for HIV which they attributed to adequate funding. In contrast, medication for NCDs was frequently out of stock which complicated multimorbidity management. Healthcare workers expressed concern over the frequent drug stockouts, considering that both facilities act as referral centres.

*“We’re doing well in HIV because we have a lot of support from organisations, but we struggle with hypertension and diabetes drugs. It’s very difficult for patients who have multimorbidity because they’ll only find one or two drugs at the pharmacy. They come back with complications not because of suboptimal drugs, but because they’re not on one of their drugs.”* (QECH, HCW08, female,4years’experience)

The lack of medication was felt to contribute to patient mistrust of health facilities.

*“For instance, some patients complain that every time they go to the hospital, they are told that the drugs are not available, and they say it’s just the same as staying at home because either way, they are not helped. They would rather take herbal medicine”* (QECH, HCW05,female, 10years’experience)

### Variable availability and knowledge of guidelines and policies between diseases and facilities

Malawi advocates for integrated care in its policies. However, we found a knowledge bias towards HIV and a lack of awareness of noncommunicable disease policies among all cadres of healthcare workers interviewed at both facilities due to what clinicians describe as limited training and dissemination. Clinicians were aware of the integrated disease management plans, but were not sure how that would work, considering that programmes are still siloed and felt the documents would not be as detailed as they should be to guide clinical management.

*“In particular I know about HIV and AIDs policies. I know there is a health policy, and honestly, I haven’t seen any relevance or direct application on the ground. I am not aware of a policy that talks about integrating HIV and diabetes per se*.” (QECH, HCW08, male, 4years’experience)

All clinicians had access to the Malawi Standard Treatment Guidelines, which they found useful in supporting decision-making, but some felt the guidelines did not cover complicated cases of multimorbidity. Some clinicians perceived the lack of clear protocols and diagnostic algorithms as a challenge to patient management.

*“If they have kidney disease and diabetes and or hypertension or liver cirrhosis, there is nothing that tells you what to do.” (*Chiradzulu, HCW20, female,14 years’ experience)

Clinicians reported that the HIV guidelines which were readily available at the facilities covered drug-drug interactions for patients with HIV+other communicable or NCD multimorbidity. This was not the case for patients living with NCD+NCD multimorbidity.

*“Another challenge is that the patient has multimorbidity and we need to find which drug will suit that patient. For example if the patient has CKD, and they are also on tenofovir(TDF) based ART regimen, we will need to switch them from the TDF to a renal friendly ART regimen. For HIV, the national guidelines have such, but for other NCDs we often use Google to see drug interactions.”* (Chiradzulu, HCW14, male, 5 years’ experience)

Some clinicians reported using international guidelines such as the British National Formulary (BNF) to check for drug-drug interactions.

### Poor multimorbidity reporting

We observed data clerks were responsible for completing patient data at both sites and for sending quarterly reports to the Ministry of Health for individual diseases. Even though clinicians reported listing all diagnoses in the patient’s health passport or file, hospital reports focus on single diseases.

Our participants reported data clerks and clinicians conducted joint review meetings to discuss the statistics. NCD data review meetings were infrequent compared to HIV review meetings.

*“Feedback is given to the district management team and the clinicians who attend the NCD clinic at review meetings. These meetings don’t happen as frequently as the HIV meetings.”* (Chiradzulu, HCW16, Male, 5years’experience)

### Recommendations to improve care

Our study clinicians individual and institutional insufficiencies as a barrier to quality multimorbidity care. They have made recommendation to improve coordination, efficiency and effectiveness of care. These have been summarised in box 1.

**Box 1:**
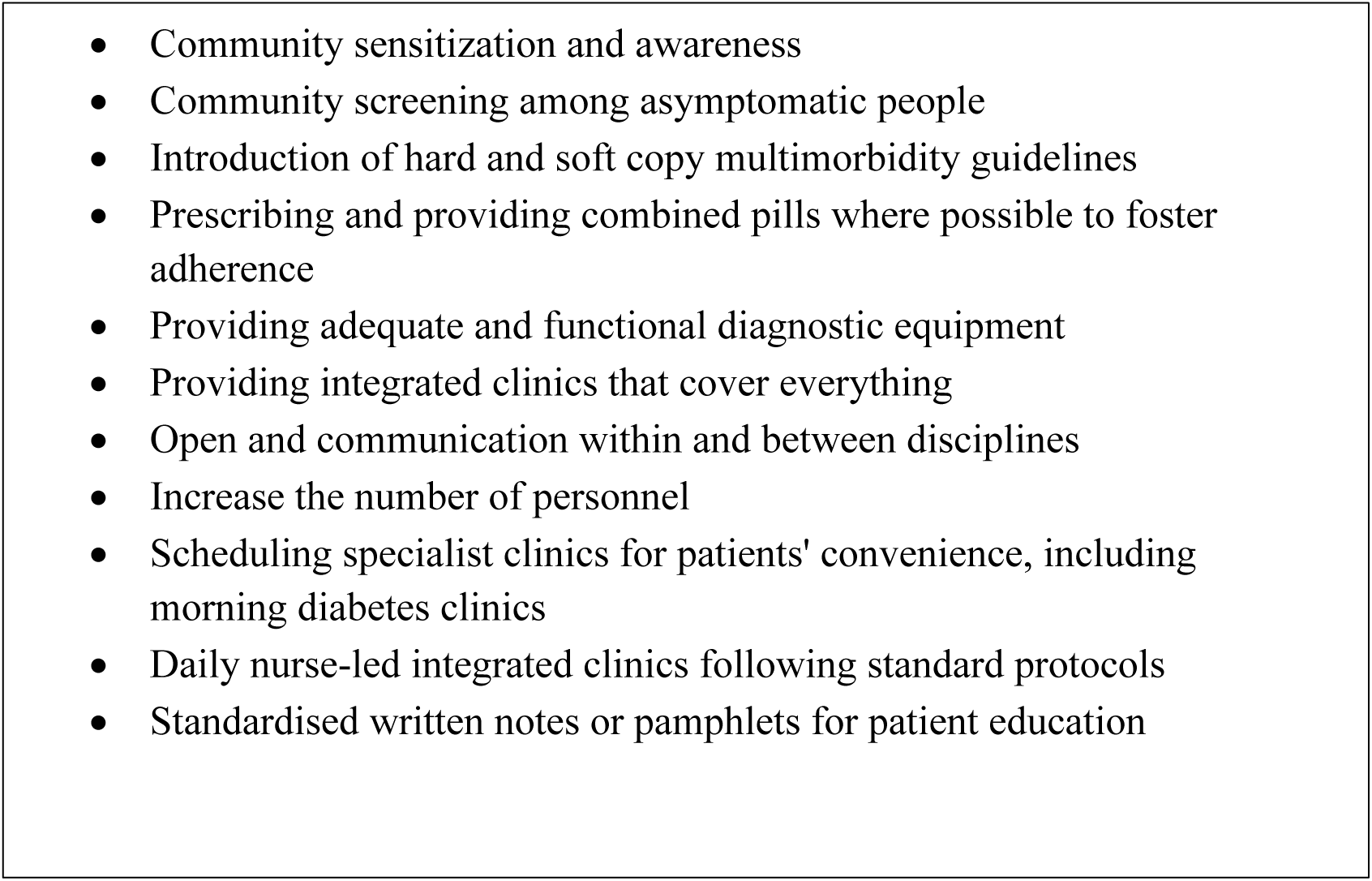
Healthcare workers’ recommendations to improve care.

## Discussion

This study reports unique insights into the perspectives of healthcare workers on how services are organised for adults living with multimorbidity presenting at hospitals in Malawi. Both facilities provide integrated multimorbidity care, but healthcare workers perceived individual (skill and knowledge) and institutional (resources) insufficiencies as barriers to quality integrated care.

Similar to findings from Tanzania and Uganda, our study participants saw HIV-NCD screening and NCD-NCD integration at Chiradzulu, as an approach for optimal management of multimorbidity(30, 31) In our setting, NCD-NCD integration only included type 2 diabetes and hypertension. Studies show that integration in sSA often falls under HIV, diabetes and hypertension.(32–35) Focusing on HIV, diabetes and hypertension as high-burden disease clusters is important. However, there is room to expand integrated care beyond these three conditions.

The desire for integration varied by level of facility. Clinicians in this study, at Chiradzulu preferred integrated clinics. This was also reported by nurses representing all levels of care in Australia.(36) Our participants and clinicians in Ethiopia preferred integrated NCD and HIV clinics as they believed this could boost the skills of the healthcare workers and make them well-rounded.(37) This is in contrast to our findings at the tertiary facility (QECH) where most clinicians believed specialised clinics are better for the patient because they offer more comprehensive care for each condition. The differences could stem from the tertiary facility having consultants and specialised clinicians wanting specialised care.(2, 38) This could mean that integration may not be a “silver bullet” to manage chronic diseases, as their complexity may still require specialised care that may not be available or appropriate at integrated clinics.

Clinicians felt they were ill-equipped and supervised to provide adequate integrated care for people living with multimorbidity. In high and low-middle income countries, healthcare workers also reported low confidence in managing multimorbidity, and highlighted the need targeted training, recruiting and retaining skilled workers for continuity of care.(36, 39, 40) It is concerning that nearly a decade after Mitambo *et al.* reported healthcare workers in Malawi are not adequately trained and supervised to provide integrated hypertension-HIV services, issues of healthcare workers’ capacity persist across various NCDs. (41)

Similar to a report from Ethiopia, clinicians in our study perceived the facilities were unable to provide adequate multimorbidity care due to limited diagnostic equipment and frequent drug stockouts.(17) Frequent drug stockouts were also reported as a barrier to care by patients living with multimorbidity in Malawi.(14, 42)While other healthcare workers reported feeling patient’s losing credibility in the eyes of the patient due to frequent drug stockouts, our findings highlight a perceived loss of credibility in the health facility.(43) Clinicians in our study felt that a lack of medication at the facility prompted patients to stay home and take unregulated herbal medicines. The perceived loss of facility credibility could result in catastrophic outcomes for patients, and spread beyond those living with multimorbidity.

Most participants reported a lack of awareness of NCD policies, which is similar to chronic respiratory diseases, with their knowledge biased towards HIV which undermined integration.(44)In a 2022 systematic review, Kassa *et al.* found NCD policies and guidelines have been poorly disseminated in sSA.(45) Maimela *et al.* argued insufficient dissemination of national guidelines for the management and control of NCDs combined with a lack of monitoring and evaluation is a key barrier to knowledge acquisition for healthcare workers.(46) Poor dissemination of NCD policies and guidelines can explain the self-reported lack of knowledge among healthcare workers. Healthcare workers in Ethiopia found the availability of clear national guidelines facilitated the implementation of HIV and NCD integration.(37)

Our findings highlight challenges in providing person-centred multimorbidity care. Other studies have also shown the importance of person-centred care, especially among people living with HIV and NCDs in the WHO African region.(47) Eswatini and South Africa deliver patient-centred care through a decentralised drug distribution for people with NCDs and HIV, and in some instances, using PEPFAR’s infrastructure.(48) According to Goldstein et al, these countries have established staff training, data systems, supply chain and policies that support the integrated dispensing of HIV and selected NCD drugs outside of hospitals and use of expert clients.

Leveraging existing HIV services to promote multimorbidity care without compromising the quality of NCD care has been feasible in this region.(49)Vice versa, integrating HIV and NCDs does not compromise HIV care or retention to care.(35) Conversely, NCDs may promote HIV services in cases of major donor withdrawal providing. opportunity for sustainable integration. Our study participants reported regularly offering counselling and health education for the patient and their carers. This can be part of the empowerment and engaging individuals and families with chronic conditions, which the WHO propose will improve outcomes through person-centred care.(5)

### Strengths and limitations

To our knowledge, this is the first paper from Malawi to report the perspectives of healthcare workers on services for adults living with multimorbidity. The complementary data collection methods and diversity of the interviewees enhanced the credibility of the data. Our study focuses on healthcare workers at two government-funded hospitals and on a narrow range of common morbidities. We did not include primary healthcare facilities, which also provide care to patients living with multimorbidity. Our findings may not apply to facilities located in various regions of Malawi or at faith-based and for-profit healthcare facilities as points of referral. Since some of the clinics reported delayed or misdiagnosis at the primary healthcare level, further studies may also explore community linkages, explore and quantify service readiness and availability for multimorbidity care.

## Conclusions

Both tertiary and secondary healthcare facilities provide care for patients with multimorbidity. However, the secondary facility lacked a strong integrated approach for individuals managing both HIV and NCDs. Health care workers perceived/reported that restricted access to resources such as diagnostic tools, medication and monitoring tools undermined their ability to manage multimorbidity. A lack of training and supervision, especially for NCDs led to limited knowledge to manage multimorbidity and provision of person-centred care. Progress from HIV services could be mirrored in other chronic diseases, and care can be provided in a decentralised manner, starting with multimorbidity screening within the communities. Facilities may model the HIV supply chain, healthcare workers’ training sessions and monitoring and evaluation tools to ensure that multimorbidity is acknowledged and sustainable. Improvements are required across all aspects of the WHO building block if facilities are to provide equitable and accessible quality care. However, Integration alone cannot solve the health system issues that this study highlighted.

## Author contributions

Conceptualisation: GTB, IS, FL, JR, RM, MT Data collection: GTB, IS

Formal data analysis: GTB supported by IS, Supervision: FL, RM, MT

Visualization: GTB Drafting manuscript: GTB

Review and editing: All authors

## Data Availability

All data generated or analysed during this study are included in this published article [and its supplementary information files]. Anonymised copies of original transcripts can be shared upon reasonable request.

## Acknowledgements

We would like to acknowledge the management and staff at our study sites for their support during data collection. We would like to thank members of the Multilink Consortium for their technical support (Sup 4).

## Supplemental Information

S1_Data Collection tools

S2_Standard Reporting for Qualitative Research Checklist S3_ Consortium Reflexivity Statement

S4_List of Multilink Consortium members

